# Evaluation of the Safe Recovery Program to reduce falls in older people in hospital: Protocol for a multicentre stepped-wedge cluster randomised trial

**DOI:** 10.64898/2026.08.26.26361288

**Authors:** Anne-Marie Hill, Meg E. Morris, Leon Flicker, Christopher Etherton-Beer, Adam Semciw, Steven M. McPhail, Catherine M. Said, Ronald I. Shorr, Caroline Bulsara, Katherine Harding, Amy T. Page, Bodil Rasmussen, Max Bulsara, Hazel Heng, Jacqueline Francis-Coad, Kerry Mace, Rebecca Woltsche, Kelly-Ann Hahn, Uyen Phan, Carol Watson, Stephen Peterson, Donald Campbell, Terry Haines

## Abstract

**Background:** Falls in hospitals are associated with injuries, deaths and poor patient outcomes. Although clinical guidelines recommend educating hospital patients about how to prevent falls, not all hospitals systematically deliver evidence-based patient falls education. The primary aim of this study is to implement and evaluate the effectiveness of delivering a research-informed education program called the “Safe Recovery Program” with ward support on rates of falls and falls-related injuries in hospitals. The secondary aims include measuring changes in patient and staff knowledge and awareness about falls prevention and identifying barriers and facilitators to staff and patients taking action to reduce hospital falls.

**Methods:** The trial will adhere to the Consolidated Standards of Reporting Trials guidelines. Twelve wards will be recruited from five Australian hospitals over a 65-week period. A stepped-wedge cluster randomised controlled trial design will be used with unidirectional crossover from control to experimental conditions together with randomisation of when each cluster makes the transition. The crossovers will occur at 12 timepoints, each five weeks apart. Alongside the trial, patients and staff on participating wards will be recruited for interviews and qualitative data analyses will be conducted to understand how to optimise implementation.

The experimental condition involves usual care plus delivery of the Safe Recovery Program. For the Safe Recovery Program, supervised allied health assistants will deliver brief falls education programs to all suitable patients in designated wards, reinforced by all ward staff. Falls champions, who are registered nurses and allied health professionals, will provide Safe Recovery Program training for staff, using a train-the-trainer model. The ward staff will also be trained in how to support hospital patients to adopt safe behaviours. The primary outcome will be falls per 1000 patient bed days. The secondary outcomes will be: (i) injurious falls per 1000 patient bed days (ii) patient and staff changes in falls awareness, knowledge and motivation; and (iii) barriers and enablers to hospital staff engaging in behaviour change and program implementation. An economic evaluation will also be conducted to estimate the incremental cost effectiveness of implementing the Safe Recovery intervention.

**Ethics and Dissemination:** Ethics approvals have been obtained from The Royal Melbourne Hospital Human Research Ethics Committee (HREC/113864/MH-2024).

The findings will be disseminated through peer-reviewed journals, workshops and conferences. Consumer team investigators will guide the communication of findings to the target audiences, including older patients, hospital staff, healthcare managers and policy makers.

**Trial Registration Number:** ACTRN12624001469505

## BACKGROUND

Falls in older people are an increasing global health burden.^1^ Up to 1 million falls occur every year in hospitals in the US alone ^2^ and over 80% of these are in older adults. Falls injuries acquired in hospital are associated with people experiencing longer lengths of stay, being discharged with pain and disability and being admitted to residential aged care.^3–8^ Between 30-50% of hospital falls cause injury, with hip fractures and head injury being the most serious.^3^ Hospital falls are also correlated with higher rates of anxiety, loss of confidence, and reduced independence that can affect quality of life.^9,10^ Older people are particularly vulnerable to falls as they often have multiple co-morbidities or limited functional reserve.^1,3,7^

Prior research has provided evidence that delivering falls prevention education for older patients whilst they are in hospital can reduce falls and fall-related injuries.^9,11–13^ Hill et al.^11^ conducted a stepped-wedge, cluster-randomised controlled trial showing that falls rates in rehabilitation units declined after individualised patient and staff education programs. A systematic review of the international literature^14^ provided corroborating evidence that patient education was an effective method for reducing hospital falls. Patient falls prevention education was the only intervention with Level Grade 1A evidence for reducing hospital falls in the 2022 World Falls Guidelines for falls prevention in older adults.^1^

Hospital falls differ from home and community falls. Research has shown that complex, unfamiliar hospital environments, limited patient-staff communication and staff actions are key contributors to hospital falls.^15,16^ Over 80% of hospital falls occur when patients are alone.^17^ Patients may attempt to move around unfamiliar hospital rooms or go to the toilet when they are unsteady or ill,^18,19^ thus increasing their risk of falls and associated injuries. Falls prevention education facilitates staff engaging directly with older patients, so they understand risk and safe behaviour in hospital and hence, is critical to reducing hospital falls^1,20–22^

However, a major gap exists because few hospitals appear to systematically implement falls preventive education.^20^ Both older adults and staff have identified barriers to delivering patient falls education in hospitals.^13,23–27^ Staff identified that barriers included patient illness, patient risk-taking behaviour, time and resource shortage and communication barriers with non-English speaking patients.^24,25^ Older patients have identified that there is insufficient falls education in hospital resulting in uncertainty about how to engage in falls prevention.^26^ Both patients and staff identified enabling strategies of timing of delivery around wellness and the patient’s mobility; tailoring messages for each older patient including from culturally diverse backgrounds; and key staff members being assigned to lead program delivery.^25–27^ Using these findings, we aim to implement a patient education program in multiple hospitals, using a ward-based approach and a hybrid effectiveness-implementation design.^28^

The overall objective of this project is to improve the outcomes of older hospital patients by implementing an individualised patient falls education program, embedded with health professional and clinician researcher training, to reduce falls. The patient education will use the revised Safe Recovery falls prevention education program which updated the original resources and was developed in partnership with older patients and hospital staff.^9,11,29^

There are four associated aims:

**Aim 1**: Co-produce resources and strategies with consumers, hospital staff, and health services which will support delivery of the Safe Recovery education program in hospitals, tailoring resources for each health service.

**Aim 2**: Evaluate the effectiveness of implementing the Safe Recovery falls prevention patient education program with ward support on rates of falls and rates of falls-related injuries.

**Aim 3:** Evaluate the impact of implementing this falls prevention patient education program led by falls champions on patient and staff falls awareness, knowledge, and engagement in falls prevention strategies and identify barriers and enablers to staff and patients taking action to reduce hospital falls.

**Aim 4**: Conduct an economic evaluation of implementing the Safe Recovery intervention to assess cost-effectiveness for health services.

## METHODS

### Ethics

Ethics approval for all sites has been obtained from The Royal Melbourne Hospital Human Research Ethics Committee (HREC/113864/MH-2024) and each site provides governance approval. The trial was prospectively registered in the Australian New Zealand Clinical Trials Registry (ACTRN12624001469505). The study complies with the Consolidated Standards of Reporting Trials (CONSORT) guidelines: extension for stepped-wedge cluster RCT.^30^

The research team gained a waiver of consent from the Ethics Committee for collecting routinely collected hospital data (effectiveness evaluation) as well as informed consent for staff, patient and family interviews (implementation evaluation) as per guidance in the National Health and Medical Research Council Australia national statement 2023.^31^

### Design

A hybrid Type II effectiveness-implementation trial (stepped-wedge cluster RCT) will be conducted. This design is useful for assessing interventions that have prior evidence of efficacy.^28,32^ The trial evaluates *clinical effectiveness* and simultaneously gathers information about *implementing* falls prevention patient education in a real-world hospital setting.^28^The stepped-wedge cluster RCT is a pragmatic design for service delivery interventions where the outcome data (for this study being falls) are routinely recorded and easily accessed.^32^ The trial commenced on the 22^nd^ September 2025 and is scheduled to be completed on the 11^th^ October 2026.

The stepped-wedge cluster RCT will use a unidirectional random, sequential cross-over from control to intervention conditions (Figure 1). The crossovers will occur at 12 different timepoints, each five weeks apart (total 65 weeks). Wards will be randomised to their start date, with one ward per step. Where local stakeholders indicate that two wards at the same hospital could be at risk of contamination due to proximity or organisation structure, they will be randomised together to mitigate risk of contamination. An independent investigative team will randomise wards to conditions.

**Figure 1.**
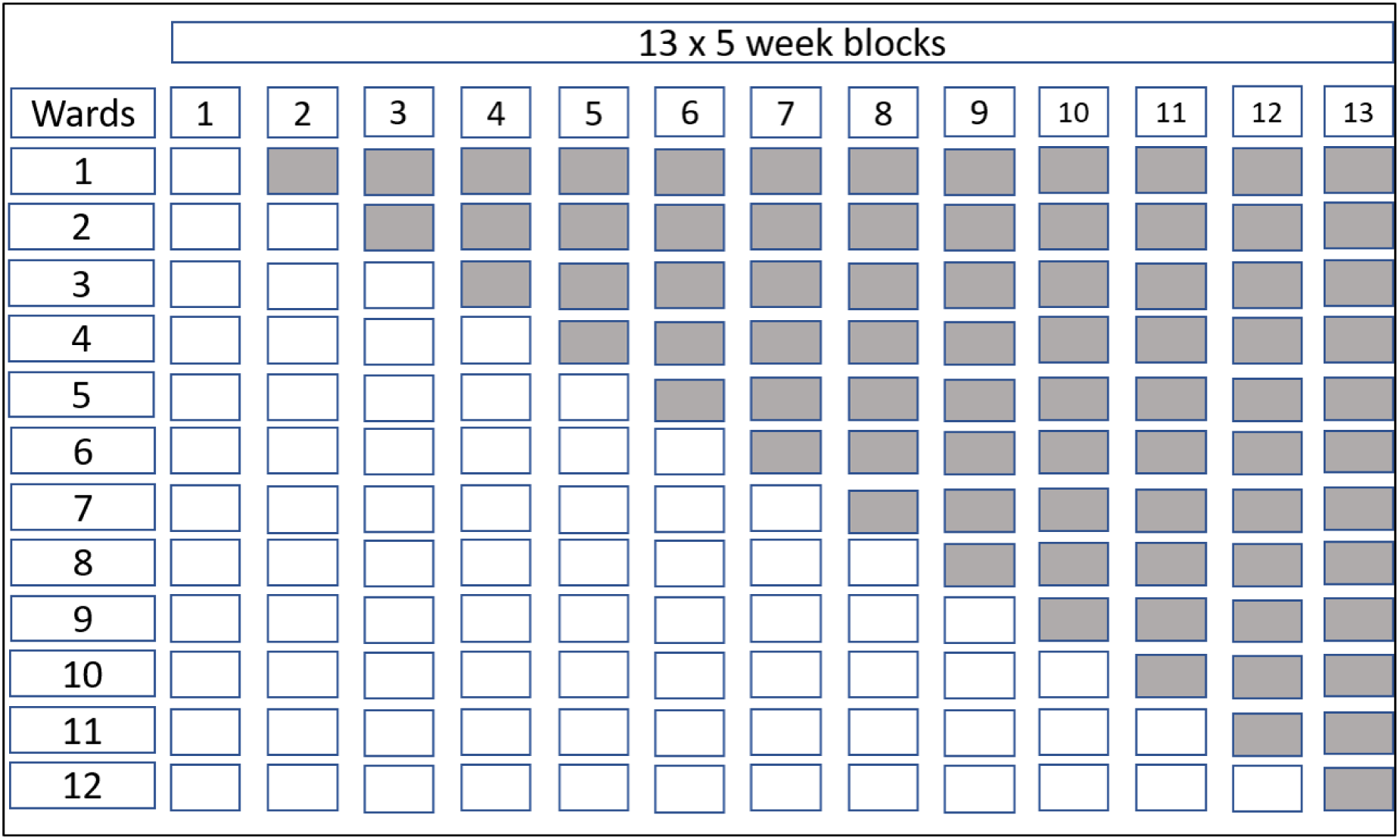
Stepped-wedge design: Shaded cells represent interventional period.

The primary effectiveness outcomes (falls) will be measured at the ward level using routinely collected hospital data. Implementation outcomes will draw upon audits of ward activity, records of intervention delivery, and semi-structured interviews with patients, families, falls champions and staff.

### Inclusion Criteria

(i) Hospital wards. The hospital wards for the trial will include medical, acute, aged care, rehabilitation and surgical wards. They will be selected from Royal Perth Hospital (Western Australia), Northern Health (two hospitals, Victoria, Australia) and Western Health (two hospitals, Victoria, Australia).

The education will be delivered on the wards to the patients who are screened by their clinical team as being sufficiently medically stable to receive the Safe Recovery Program. For patients to receive the education they need to be ≥ 18 years of age, predicted to be on the ward at least 48 hours, have the ability to engage with the allied health assistant or allied health professional (e.g. medically stable at time of interaction) and have normal cognition or only mild cognitive impairment. Exclusion of an individual patient for cognition by the treating team would be considered if the 4 A’s delirium test (4AT) ≥4/12,^33^ Abbreviated Mental Test Score (AMTS) p<7/10,^34^ or Shortened Abbreviated Mental Test Score (ATM) > 4/12,^35^ with final judgement made by the patient’s medical team. Patients who have acute confusion /delirium will be monitored by the treating team for resolving delirium and the Safe Recovery Program will be delivered if their delirium resolves. Family members /carers will be included when on the ward by discussing the falls prevention strategies with them and they can view the resources with family. Patients who do not speak English will receive education if their family are present on the ward to assist or if interpreting services can be provided.

ii) Individual patients and staff (interviews for implementation outcomes). Patients on the ward who can provide written informed consent will be eligible to be interviewed about the program. Staff older than 18 years of age who work on participating wards will be eligible to be included in the trial and interviewed about their experiences on the ward. Staff or patients who receive an education session/ resource will be eligible to complete a de-identified online survey to give feedback.

### Exclusion Criteria

i) Hospital wards that provide care for children <18 years, ante or post-natal services, intensive care, high dependency units, short stay ambulatory units or emergency departments will be excluded. These wards have a different context for falls events and therefore differing tailored strategies are required. Surgical units will only be included if the patient population includes large numbers of older adults admitted and if the average length of stay is greater than 48 hours.

ii) Individual patients and staff (implementation outcomes). Any patients who are deemed too unwell, or unable to be approached for other reasons (e.g. infection control, acute confusion) will be excluded by ward staff from the patient education component of the intervention and will also be excluded from participating in an interview. Where patients have diagnosed dementia or acute confusion, their family members may be asked to participate in an interview to seek their reflections on the patient’s experience on the ward. Staff who have not worked on participating wards for at least six weeks will be excluded from being interviewed about the program as they will have limited knowledge and experience of the Safe Recovery intervention.

### Recruitment

i) Ward level (effectiveness outcomes). Hospitals lead site investigators at the participating hospitals will invite hospital managers, nurse unit managers, allied health staff and quality and safety staff to express interest in their wards participating in the trial. Wards for the trial will be selected based on the inclusion criteria above. A waiver of consent has been obtained from the ethics committees for evaluating the effectiveness of the intervention, as the primary outcome of falls will be evaluating using de-identified hospital data. All patients who receive the intervention will be invited to complete a short questionnaire, using a de-identified survey link, with consent implied by survey submission.

(ii) Individual hospital patients (implementation outcomes). Patients receiving care on participating wards and/ or their family members will be invited to participate in interviews. Eligible patients will be identified by ward staff, who will ask for verbal permission for researchers to contact them to invite them for an interview. The ward staff will approach patients who have no acute medical issues or moderate-severe cognitive impairment precluding participation. When verbal permission is given by a patient, a trained site researcher will approach the patient and/or their family in-person or electronically by phone or email to discuss the study and go over the patient and /or family participant information and consent form (PICF). Patients who do not speak English will be considered for interview if present on the ward. Staff will seek verbal permission from family for researchers to approach these patients, with their family present. A trained site researcher who speaks the patient’s language or an interpreter will assist in conducting the interview if the patient agrees to participate. A purposive approach will be used with stratification of the sample to be employed at each site. The site researcher will monitor recruitment to seek diversity of patient perspectives on the ward based on patients’ age, gender, cultural background and falls risk as recorded in their clinical care.

(iii) Individual hospital staff (implementation outcomes). Staff who work on participating wards in the trial will be identified by the nurse unit manager who will seek verbal permission from the staff member for researchers to contact them to invite them for an interview. The site researchers will provide the nurse unit manager with information about how to select a broad range of potential staff participants that meet the purposive inclusion criteria. When verbal permission is given, a trained site researcher will then approach the staff member in-person or electronically by phone or email to provide information about the study and review the staff PICF. A purposive approach will be used with stratification of the sample to be employed at each site. The site researcher will monitor recruitment to seek diversity of staff perspectives on the ward based on gender, cultural background and years of experience working in a hospital setting.

Online surveys after any education that is delivered (implementation outcomes) will provide an opt-in check box for use by ward staff, train the trainer staff, and by patients who receive an individual education session / resource. Participants can complete or choose to not complete these de-identified surveys.

### Randomisation

Codes for each hospital ward will be created by the chief investigator (AMH) who is not involved in any site and used to allow for blinded randomisation of the 12 wards, using a stratified approach for the three sites to ensure each site is able to commence delivery on their wards at staggered periods. The randomisation will occur before the start of the trial using a computer-generated number sequence by an external team led by an investigator (SM) who is not associated with the chief investigator’s university, any of the hospital sites, recruitment, intervention delivery or trial procedure. The results will be communicated to the Chief Investigator (AMH) and the three Site Investigators (CEB-Royal Perth hospital, Western Australia, AS-Northern Health, Victoria and CMS-Western Health, Victoria) only. Each individual ward will be informed of their allocation by the Chief and Site Investigators eight weeks prior to their step into the intervention, to allow time for the champions and ward to undertake training and resource preparations for entry

### Blinding

It is not possible to blind ward staff intervention providers to their role in the trial as they need to undergo training and deliver the intervention. Individual recruitment of participants does not occur and hence most patients are likely to remain unaware the trial is in progress and not observe that a new education program is being delivered. All data sent to the chief investigator will be de-identified and data analysts will be blinded to treatment conditions.

### Interventions

#### Experimental Intervention

The intervention is an individualised patient falls prevention program (the revised Safe Recovery Program)^29^ which will be provided to eligible hospital patients in the participating wards with staff training and support in addition to usual hospital care. The Safe Recovery Program education includes the message “*3 simple steps for stopping falls”:* i) Know if you need help to get up and walk around; ii) Ask for help; iii) Wait for help.^9,29^ The Safe Recovery Program will provide patients with personalised falls prevention education, goal setting to prevent falls and falls information, reinforced by ward staff. The implementation has six main elements:

1. ward staff training and ongoing program leadership throughout the trial led by trained “falls champions” who are trained senior allied health or nursing professionals (2 per site).
2. a multimedia education package (consists of the Safe Recovery patient guide, Safe Recovery brochure, a Safe Recovery patient plan and a choice of two videos (3 minute and 8-minute versions, with guides, brochures, and videos available in English, Mandarin, Vietnamese and Arabic).
3. initial 5 to 10 minutes conversations with a patient delivered by a trained allied health assistant (supervised by allied health professionals and nursing staff) who will assist the patient to develop a short, written plan to prevent falling in hospital.
4. recording of education delivery in the patient care plan and follow up and reinforcement of falls prevention education by ward staff as part of delivering care.
5. further individual discussions with patients by allied health assistants or allied health professionals as required, as part of usual care to continue to tailor plan.
6. training of hospital ward staff throughout the trial using a train-the-trainer model. Ward staff will be regularly encouraged by falls champions to embed strategies (e.g. place call bell in reach of patient, ensure all patients have walking aid, inform family, include falls information in handover) into care for all patients on the ward. Brochures and ward posters will be used to reinforce messages to all staff and patients.

The initial short discussion will occur with the trained allied health assistant who will be supervised by allied health professional or nursing staff. All allied health assistants will receive a training workshop prior to commencing education delivery. The discussion alerts the patients that they personally are at risk of falls during admission, gives them knowledge about the nature of falls, as well as knowledge about the benefit of engaging in falls prevention activities and, explains cues to action and facilitates self-efficacy to take action. The subsequent discussion sessions by all staff will support the patient in building upon the knowledge to prevent falls and to review the patients’ goals as their mobility changes over time. The patient plan consists of two to three short, easily remembered messages (e.g. use my frame to walk) and the allied health assistant will encourage the patient to write their plan to remain safe. As part of the program, patients will be encouraged to identify their awareness, knowledge and motivation pre and post education via a short de-identified survey administered by the allied health assistant or the therapist.

### Control Intervention

The control condition will be usual hospital care, which includes falls risk management for all patients on participating wards and implementation of organisation-wide safety and quality policies and programs.

### Outcomes

#### Primary Outcome (effectiveness)

The primary outcome will be falls per 1000 patient bed days. Patient falls that occur during hospitalisation on a unit involved in the trial are defined as ‘an event which results in a person coming to rest inadvertently on the ground or floor or other lower level’.^1^ Falls rates for each ward will be measured by using the number of falls that occur during the time of the trial alongside the total number of hospital bed days used by patients (patient length of stay) on participating wards. Falls and number of bed days used in a ward are routinely collected hospital data.

#### Secondary Outcome (effectiveness)

Injurious falls per 1000 patient bed days. An injurious fall is defined as any fall recorded as causing an injury in the sites’ adverse event reporting system. Injurious falls rates will be measured using the number of injurious falls that occur during the time of the trial alongside the total number of hospital bed days for patients (patient length of stay) on participating wards. Information about all injurious falls is routinely collected hospital data.

#### Secondary Outcomes (implementation)

Implementation outcomes have been based on Consolidated Framework for Implementation Research (CFIR) domains.^36^ Outcomes will be patient and staff (including falls champions and trainers) changes in falls awareness, knowledge and motivation, measured using online baseline and post-intervention surveys. Ward engagement in falls prevention strategies will be measured using audits of ward-level data (e.g., handover records) / bedside charts (e.g. patient goals). Intervention reach, adoption, implementation and sustainability will also be measured, including the proportion of patients/wards who receive the education, and number of staff training sessions. Semi-structured interviews with falls champions, patients, families, and staff will identify barriers and enablers to behaviour change and program implementation at the individual, ward and organisational levels.^36^

##### Healthcare resource use and costs

Health care resource use will be collected from the perspective of a health service deciding whether or not to implement the Safe Recovery program intervention in comparison to their usual care. This will include healthcare resource use and costs routinely recorded via hospital administration systems, including post-fall-related usual care whilst in hospital. We will also record trial-related intervention resources use (e.g., staff training, resources for delivery of the Safe Recovery Program by staff). Healthcare resource use and costs will be used to inform a trial-informed economic evaluation which will be reported separately.

#### Sample Size

The trial will involve 12 wards across two Australian states: Royal Perth Hospital Western Australia (1 hospital, 4 wards), Northern Health Victoria (2 hospitals, 4 wards), and Western Health Victoria (2 hospitals, 4 wards), which collectively provide over 2000 beds and admit large numbers of older patients. With admission rates that are high and consistent, these wards are projected to include approximately 6000 to 9000 admissions over a 12-month period (approximately 2500 admissions at Royal Perth Hospital, 3500 across Northern Health, and over 3000 across Western Health). The trial will span 65 weeks, with patient admissions across these settings providing a robust sample size to adequately power the study.

#### Power Calculations

Power calculations for the primary effectiveness outcome (falls rate) used Monte Carlo simulations of the proposed trial design. These indicates that the study will achieve 80% power to detect a falls rate reduction from 6.2 to 4.2 falls per 1000 patient-days (32% reduction), using a 2-tailed alpha level of 0.05, assuming a standard deviation of 2.4 falls/1000 patient-days and a low intra-cluster correlation (ICC < 0.0001). These power analysis inputs were based on data collected over 47 ward-months across the hospital trial sites.

### Statistical Analysis

#### (i) Statistical analysis (effectiveness outcomes)

Fall rates and injury rates, will be compared using generalised linear mixed models following recommended stepped-wedge cluster RCT model specification.^37^ This includes a random cluster effect and random cluster by step effect. Each step period will be modelled as a fixed categorical effect to account for potential secular trends. A Poisson or negative binomial distributional family for the outcome will be considered and selected based on data fit. The effect variable will switch from 0 to 1 as each cluster changes from control to intervention steps. Unadjusted results (except clustering) will be reported. Sensitivity analysis will consider a potential linear intervention effect to account for increasing familiarity with the intervention. Outcomes will be expressed both as Incidence Rate Ratio and mean difference in rate of falls outcomes/1000 occupied bed days.

#### (ii) Statistical Analysis (implementation outcomes)

##### Quantitative data

Intervention delivery measured from research records, including proportion of patients who receive education, number of patient and staff training sessions, and associated surveys, will be summarised using descriptive statistics, then analysed as appropriate to evaluate implementation fidelity, changes in awareness, knowledge and engagement in falls preventive strategies.

##### Qualitative data

Data from interviews and open-ended responses from surveys will be analysed. All data will be de-identified to protect participant confidentiality. Themes will be developed inductively, based on the data itself, or deductively, informed by the guiding theoretical framework (CFIR).^36^ Multiple researchers will review and discuss themes to enhance rigor, minimise bias, and ensure that interpretations are grounded in participants’ experiences. NVivo software (Lumivero, version 15, 2025) will be used to assist with coding and organising the data systematically. Findings from all datasets will be merged as a matrix to compare ward, hospital and organisation level implementation barriers and enablers to success using the CFIR framework.

##### Healthcare resource use and costs

Healthcare resource use and costs collected from this trial will be used to inform cost-effectiveness modelling to estimate the incremental cost (healthcare system perspective) of implementing the intervention per fall averted. This will include trial-related intervention resource use (e.g., staff training, delivering Safe Recovery Program) and other healthcare use associated with usual care including post-fall care.

### Handling of Withdrawals

For the stepped-wedge trial, there will be no withdrawals at the patient level, as a waiver of consent will permit data capture for all participating wards. However, for the process evaluations individual participants (patients or staff) may withdraw from qualitative interviews for any reason after consenting, prior to analysis. If they withdraw after the interview has already been conducted, their contributions will be identified and immediately deleted from the interview recordings and transcripts. Participants will be informed, prior to giving consent, that once transcripts are de-identified and analysis has commenced, exclusion of data will no longer be possible.

### Risk Management and Safety

The research team operating through the trial steering and governance committees, comprise senior leaders from all participating hospitals. The trial will be conducted according to approved ethics and governance protocols.

Falls occur frequently on hospital wards, with more falls known to be reported on medical wards and wards that admit large numbers of older patients.^38^ Patient falls are known to occur on all wards participating in the trial and mandated hospital policies and procedures apply for providing falls risk assessment and management commencing on admission and post-falls treatment and care. These policies and procedures will be applied for any patient who falls on participating wards, regardless of whether they received any component of the intervention.

Falls rates will be continuously recorded and monitored on participating wards through hospital safety and quality procedures. The intervention is low risk, involving wards delivering a new means of patient education embedded within the existing education and care already provided by the ward staff. As with any other care on the ward, patients can immediately inform their medical team if the education or participating in an interview in any way causes distress (e.g. patient has a strong fear of falls and does not want to discuss falls safety) and the team can provide appropriate support.

### Data Management, Data Security and Data Handling

All data will be collected and stored in adherence to the ethics approvals, which include the approved data management plan, and governance approvals of the three sites.

#### Hospital Routinely Collected Data (Effectiveness and Economic Analysis)

Only de-identified electronic data will be transferred to The University of Western Australia (UWA) secure storage. De-identified electronic data will be transferred directly from the Business Intelligence Unit from the three sites to UWA secure storage using MYFT for WA Health data^39^ and AARNet FileSender^40^ for Victorian sites, which allows universities to send and receive sensitive data using encrypted transfer. Files will be encrypted during transfer, and the Site Investigator will set an expiry date and provide a download notification to further control the security of the data. Data are accessed only by approved members of the research team.

#### Implementation Outcomes Data

Participant informed consent for interviews will be obtained via the REDCap digital online database at each site or with a hard copy consent form if online REDCap is not available on the ward. and automatically stored in the hospital secure folder. Process evaluation data (patient and staff interviews, surveys, ward audits and allied health records of education delivery) will be stored digitally in initially in re-identified format with use of a code and subsequently in de-identified form during the project data collection phase at the sites in their secure restricted servers. These data will be stored at the hospital sites, following their governance requirements and approved data management plan, until secure transfer to UWA secure storage at conclusion of data collection.

#### Data Confidentiality and Security

No personally identifiable data will be collected as a part of this study. Hospital records for hospital falls data and patient admissions to wards, such as length of stay data will be collected without any identifiable or re-identifiable information about individual patients. These data will also be analysed and reported in an aggregated manner. Participants will receive a plain language summary of the project. This will contain no identifying information about individual patients.

Qualitative data from interviews will be de-identified at sites after data are checked for completeness, replaced by a participant ID number and transferred to UWA for secure storage, with access only via the data custodian, Chief Investigator (AMH). Data will be analysed and grouped across all interviewees, with no attributable data linked to individual staff or patients in final reports and publications. Online surveys will be de-identified at the time of data collection with no personal information collected. All data stored at the sites until final transfer to UWA secure storage are online in restricted folders with password and staff identified access only available to the Site Investigator. Where necessary for data collection or management, named CI/ AI investigators on this application and project staff at each site will receive authorised, monitored access from the Site Investigator.

Data will be stored securely at UWA and subject to the Western Australia University Sector Disposal Authority (WAUSDA) for retention and disposal. In accordance with this authority, the datasets and original ethics applications will be retained securely for 15 years after date of publication of the results, or 15 years after conclusion of the project, whichever is later, then destroyed.

#### Adverse Events

Site investigators (CEB, AS, CMS) monitor the project and respond to any adverse events. Any adverse events perceived to be related to the trial will be reported to the Site Investigator, who will report to the Chief Investigator (AMH). Any adverse events will be immediately reported to the approving HREC, local site governance, and the steering committee and amendments to the trial protocol made if required.

#### Data Monitoring Committee

A Data Safety Monitoring Committee (DSMC) has been implemented. The DSMC consists of senior investigators who are not involved in site management or intervention delivery (AMH, MEM, MB, SM, TH). The chief investigator informs the committee about the progress of the trial at each step into the cluster and after each steering committee meeting (three monthly). The committee can be convened if any unanticipated events occur.

### Consumer Engagement

Consumer involvement will be assured through co-design and consumer participation throughout the project. Representing consumer interests, two consumer representatives, one in Western Australia (KM) and another in Victoria (SP) are named as associate investigators on the team. Consumer engagement meetings will be held four times yearly. The consumers will be invited to review and contribute to each stage of the clinical trial, up to and including supporting the dissemination of research findings attributed to this trial. They will provide feedback and input when the online staff education training resources are designed, for example, being asked to interact with draft resources and view online links to the education. Ongoing tailoring and final co-production of supportive resources for the education was initially achieved through two consumer workshops (one in Western Australia and one in Victoria) which have already been completed. The consumers at the workshops comprised of older people and their family carers living in the Royal Perth Hospital (Western Australia), Northern and Western Health (Victoria) catchment areas in Australia.

## DISCUSSION

This study will provide evidence for how a patient education program that has been found to significantly reduce hospital falls^9,11^ can be more widely implemented across diverse wards and hospitals. The study has potential to improve the effectiveness of health services by improving safety, quality and efficiency through reducing hospital falls. Australian hospitals spend an estimated AUD$590 million yearly in resources trying to prevent falls, yet few hospital falls prevention programs are evidence based.^41^ A Cochrane review that synthesised evidence for falls prevention in hospital reported that tailored patient education probably reduces the rate of falls and risk of falling.^42^ However, a global analysis of falls guidelines found that barriers exist to implementation of evidence into clinical practice, including ambiguities in how staff and patient falls education should be conducted.^20^ Hence, systematic implementation of patient education has been limited. These implementation gaps were exemplified during a survey of 24 hospitals in the United States that was conducted to identify falls prevention implementation strategies being used in hospitals. The findings identified an overreliance on less time-intensive universal practices, rather than tailored patient falls prevention strategies, as well as few implementation strategies to enhance staff adherence to falls prevention practices.^43^

The study is expected to benefit broader hospital and consumer groups after it concludes because we are embedding systems for informing consumers about falls prevention and training and capacity building for reducing falls for health professional staff and hospitals. All resources will be made freely available to Australian hospitals after the project, to more extensively translate falls prevention evidence to practice.

## Data Availability

Due to hospital restrictions, data are not available to share.

## Access to Data

Due to hospital restrictions, data are not available to share.

## Author Contributions

*Conceptualisation,* AMH, MEM, LF, CEB, AS, SM, CMS, RS, CB, KH, AP, BR, MB, HH, JFC, SP, TH

*Formal analysis,* AMH, MB, SM, TH, AS, CEB

*Funding acquisition,* AMH, MEM, LF, CEB, AS, SM, CMS, RS, CB, KH, AP, BR, MB, HH, JFC, KM, RW, KH, UP, CW, SP, DC

*Investigation,* AMH, MEM, CEB, AS, CMS, BR, HH, JFC, RW, UP, CW, DC, KM, SP *Methodology,* AMH, MEM, LF, CEB, AS, SM, CMS, RS, CB, KH, BR, MB, JFC, KM, SP, UP, CW, TH

*Project administration,* AMH, MEM, CEB, AS, CMS

*Supervision,* AMH, MEM, CEB, AS, CMS, DC, RW, KH, UP, CW, DC

*Writing,* original draft, MM, AMH, TH

*Review and editing*, All authors have reviewed and agreed to the final submitted version of the manuscript.

